# Intensity and longevity of SARS-CoV-2 vaccination response and efficacy of adjusted vaccination schedules in patients with immune-mediated inflammatory disease

**DOI:** 10.1101/2022.04.11.22273729

**Authors:** David Simon, Koray Tascilar, Filippo Fagni, Arnd Kleyer, Gerhard Krönke, Christine Meder, Peter Dietrich, Till Orlemann, Johanna Mößner, Julia Taubmann, Melek Yalcin Mutlu, Johannes Knitza, Stephan Kemenes, Anna-Maria Liphardt, Verena Schönau, Daniela Bohr, Louis Schuster, Fabian Hartmann, Moritz Leppkes, Andreas Ramming, Milena Pachowsky, Florian Schuch, Monika Ronneberger, Stefan Kleinert, Axel J. Hueber, Karin Manger, Bernhard Manger, Raja Atreya, Carola Berking, Michael Sticherling, Markus F. Neurath, Georg Schett

## Abstract

**Objectives:** To investigate the intensity and longevity of SARS-CoV-2 vaccination response in patients with immune-mediated inflammatory disease (IMID) by diagnosis, treatment and adapted vaccination schedules.

**Methods:** SARS-CoV-2 IgG antibody response after SARS-CoV-2 vaccination was measured longitudinally in a large prospective cohort of healthy controls (HC) and IMID patients between December 2020 and 2021. Demographic and disease-specific data were recorded. Humoral response was compared across treatment and disease groups, and with respect to receipt of booster vaccinations. Age and sex adjusted SARS-CoV-2 antibody response was modelled over time. Marginal mean antibody levels and marginal risks of poor response were calculated at weekly intervals starting from week-8 after the first vaccination up to week 40.

**Results:** Among 5076 individuals registered, 2535 IMID patients and 1198 HC were eligible for this analysis. Mean antibody levels were higher in HC compared to IMIDs at all-time points, with peak antibody response in HC more than twice that in IMIDs (12.48 (11.52-13.52) vs. 5.71 (5.46-5.97)). Poor response to vaccination was observed in IMID patients treated with agents affecting B- and T-cell functions. Mean differences in antibody response between IMID diseases were small. After additional vaccinations, IMID patients could achieve higher antibody levels than HC vaccinated according to the two-dose schedule, even-though initial antibody levels were lower.

**Conclusions:** IMID patients show a lower and less durable SARS-CoV-2 vaccination response and are at risk to lose humoral immune protection. Adjusted vaccination schedules with earlier boosters and/or more frequent re-doses could better protect IMID patients.

## Introduction

Severe acute respiratory syndrome coronavirus 2 (SARS-CoV-2) constitutes a significant threat to patients with immune-mediated inflammatory diseases (IMIDs). Due to the immune dysfunction and use of immune-modulatory drugs, hosts responses to vaccination is altered in IMID patients and can vary considerably in terms of efficacy (1, 2), longevity (3), and protection against poor outcome (4-6). From an immunological perspective, the majority of IMID patients are able to mount humoral immune responses after SARS-CoV-2 infection (7, 8) and vaccination (1, 2). However, these responses appear to be blunted (1, 2) and the overall prevalence of anti-SARS-COV-2 antibody positivity due to infection is significantly lower compared to the general population (9). This seems to be at least in part driven by specific immunomodulatory therapies such as methotrexate (10), mycophenolate (11), or rituximab (12).

As reports of waning vaccine efficacy and new virus variants resistant to antibody neutralization are emerging, increased susceptibility of IMID patients for earlier breakthrough infections are of particular concern (13). To date, the immunogenicity of SARS-CoV-2 vaccines in IMID patients has been studied mostly in the first two to six weeks after vaccination, whereas extended longitudinal data are scarce. One report suggested a more pronounced decline of humoral response to vaccination in IMID patients than in healthy subjects (14). Considering that the levels of anti-SARS-CoV-2 IgG correlate with COVID-19 risk and vaccine efficacy (15), this may represent a warning signal for waning immunity, especially in high-risk subsets of patients. Therefore, the question arises whether IMID patients could benefit from adjusted vaccination schedules, with more frequent booster vaccinations.

To investigate the impact of treatment and vaccination strategies on longevity of humoral response in IMID patients and to provide evidence for current vaccination regimens of IMID patients, we analyzed a large prospective cohort of IMID patients and control subjects from the time on after the first SARS-CoV-2 vaccination.

## Methods

### Setting and Cohort

IMID patients and healthy controls (HC) were recruited from the prospective COVID-19 study program conducted by the Deutsche Zentrum fuer Immuntherapie, which since February 2020 has collected data on respiratory infections including COVID-19 as well as anti-SARS-CoV-2 antibody responses before and after the vaccination era and exposure risk behavior over time. Further study details have been described elsewhere (11). Briefly, patients attending or admitted to affiliated centers who received either no treatment or treatment with immunomodulatory agents were recruited. HC included employees of the University Hospital Erlangen, and community-dwelling individuals recruited in several organized field campaigns during the pandemic (February/April 2020; December 2020/January 2021; November/December 2021). IMID patients were asked to participate during their routine follow-up visits. A structured questionnaire was used to collect data on demographic characteristics and comorbidities. The study was approved by the Institutional Review Board of Erlangen University Hospital (#157_20 B), and written informed consent was obtained from all study participants.

### Inclusion criteria

Cohort participants with physician-diagnosed IMIDs including systemic autoimmune diseases, rheumatoid arthritis, spondyloarthritides (including psoriatic arthritis), vasculitides (excluding cutaneous-limited vasculitis), psoriasis, inflammatory bowel diseases (Crohn’s disease and ulcerative colitis), polymyalgia rheumatica and miscellaneous IMIDs including sarcoidosis, IgG-4 related fibrosing disease, juvenile idiopathic arthritis, myositis, autoinflammatory syndromes, inflammatory demyelinating diseases and control participants with no diagnosis of IMIDs were eligible for this analysis. Participants who were sampled while being evaluated for a possible IMID but remained without a clear diagnosis, those with organ specific autoimmunity (autoimmune thyroiditis, autoimmune liver disease), atopy (urticaria, asthma, atopic dermatitis), immunodeficiency, malignancy and those with uncertain diagnostic status were excluded. Included participants should have had at least one blood sample available starting from 4 weeks before their first vaccination date.

### Follow-up

Follow-up started at the time of first vaccination and continued until 01.12.2021. Samples were collected during routine visits for IMID patients, in response to email reminders for participating healthcare personnel and via several field campaigns by investigators for community-dwelling HC throughout the post-vaccination period.

### Treatment classification

We categorized current immunomodulatory treatments used for IMIDs at each sampling timepoint as cytokine- (TNF-alpha, IL-1, IL-5, IL-6, IL-12/23, IL-17, IL-36), signaling- (JAK, phosphodiesterase), adhesion molecule- (integrin), T cell- (CD80/86, calcineurin), and B cell- (CD-20,BLyS) inhibitors, conventional immune modulators (methotrexate, azathioprine, leflunomide, mycophenolate, cyclophosphamide) and other drugs (e.g. hydroxychloroquine, sulfasalazine) as detailed in Suppl.Table-1. When a patient was on multiple concomitant treatments, this was indicated as a combination treatment in a separate variable and the assignment followed a hierarchy of priority such that: biological agents> signaling inhibitors > conventional immune modulators > other drugs > glucocorticoids.

### Anti-SARS-CoV-2 antibody testing

IgG antibodies against the S1 domain of the spike protein of SARS-CoV-2 were tested by the recent CE version (April 2020) of the commercial enzyme–linked immunosorbent assay from Euroimmun (Lübeck, Germany) using the EUROIMMUN Analyzer I platform according to the manufacturer’s instructions. Optical density (OD) was read at 450 nm with reference wavelength at 630 nm. A density value of between ≥0.8 and <1.1 (OD450 nm) was considered as borderline and a value ≥1.1 was considered positive. Assays were performed in line with the guidelines of the German Medical Association (RiliBAK) with stipulated internal and external quality controls.

### Statistics

We used descriptive statistics to summarize cohort characteristics. Optical density values representing antibody levels were binned into 2- to 8-week intervals of sample acquisition time after the first vaccination for description and summarized as geometric means and standard deviations as observed.

The time course of SARS-CoV2 antibody levels were analyzed after log-transformation and modelled as a function of time after the first vaccination using mixed effects linear regression. We also modelled marginal risks of poor response using mixed-effects logistic regression, where OD values <1.1 indicated poor response. As the relationship between time and antibody levels is expected to be curvilinear, we used restricted cubic splines with four degrees of freedom for time in all analyses. All models included age and sex at the time of first vaccination for adjustment, interaction terms between time and grouping variables and participant id as a random effect. We constructed three linear models to compare i) HC vs. all IMID patients indicated by a binary variable, ii) HC and IMID diagnoses indicated by a categorical variable, iii) HC and dominant IMID treatments classified by primary mode of action. Using these models, we estimated age and sex-adjusted marginal geometric mean antibody levels at weekly intervals starting from week-8 after the first vaccination up to week 40. We performed pairwise between-group comparisons at the time of peak antibody response and at week 40 and calculated adjusted mean differences with 95% confidence intervals (CI) corrected for multiplicity by Tukey’s method. All analyses were conducted using R v 4.1.1 (R Foundation for Statistical Computing, Vienna, Austria) using packages *lme4* and *emmeans*.

## Results

### Description of the cohort

Among 5076 individuals registered in our cohort, 3733 participants were eligible for this analysis including 2535 IMID patients and 1198 HC who provided overall 5564 samples between 15/12/2020 and 01/12/2021. A flow chart describing cohort assembly is depicted in **Figure-S1**. The mean (SD) age of the cohort at the time of first vaccination was 50.4 (16.1) years and 2048 (55%) participants were females. IMID patients were older than HC, with a mean age of 55.0 (15.2) vs. 40.7 (13.5) years and more frequently female (1494 [59.1%] vs 554 [46.2%] respectively). Most common diagnoses among patients with IMIDs in order of frequency were spondyloarthritis (including PsA), rheumatoid arthritis, systemic autoimmune diseases (including SLE, systemic sclerosis and primary Sjogren’s syndrome), inflammatory bowel diseases, vasculitis and psoriasis. Distribution of diagnoses and current treatments at the time of sampling are summarized in **Table-1**. Numbers and types of vaccines administered are summarized in detail in **Suppl. Table-2**.

**Table 1.** Participant characteristics

|  | Group |  |  |
| --- | --- | --- | --- |
|  | IMID<br>n=2535 | Healthy controls<br>n=1198 | All<br>n=3733 |
| Age, mean (SD) | 55.0 (15.2) | 40.7 (13.5) | 50.4 (16.1) |
| Follow-up duration, weeks |  |  |  |
| Median (IQR) | 19.9 (12.6-27.0) | 31.1 (24.0-36.7) | 23.3 (13.9-30.9) |
| Range | 1.7-46.9 | 1.6-47.3 | 1.6-47.3 |
| Sex, n (%) |  |  |  |
| Male | 1040 (41.0) | 644 (53.8) | 1684 (45.1) |
| Female | 1494 (58.9) | 554 (46.2) | 2048 (54.9) |
| Vaccine doses, n (%) |  |  |  |
| 1 dose | 155 (6.1) | 57 (4.8) | 212 (5.7) |
| 2 doses | 2233 (88.1) | 1047 (87.4) | 3280 (87.9) |
| 3 doses | 147 (5.8) | 94 (7.8) | 241 (6.5) |
| Vaccination times, weeks, median (IQR) |  |  |  |
| First to second dose | 6.0 (5.0-6.1) | 4.7 (3.0-6.0) | 6.0 (3.9-6.0) |
| Second to third dose | 20.3 (12.0-26.7) | 29.6 (26.9-36.1) | 26.3 (16.2-31.8) |
| Number of samples per participant, n (%) |  |  |  |
| 1 | 1573 (62.1) | 741 (61.9) | 2314 (62.0) |
| 2 | 767 (30.3) | 327 (27.3) | 1094 (29.3) |
| 3 | 143 (5.6) | 111 (9.3) | 254 (6.8) |
| >3 | 52 (2.1) | 19 (1.6) | 71 (1.9) |
| Diagnosis, n (%) |  |  |  |
| Spondyloarthritis | 824 (32.5) |  |  |
| Rheumatoid arthritis | 581 (22.9) |  |  |
| Systemic autoimmune diseases | 307 (12.1) |  |  |
| Inflammatory bowel diseases | 241 (9.5) |  |  |
| Vasculitis | 184 (7.3) |  |  |
| Psoriasis | 119 (4.7) |  |  |
| Polymyalgia rheumatica | 103 (4.1) |  |  |
| Sarcoidosis<sup>§</sup> | 53 (2.1) |  |  |
| Autoinflammatory syndromes<sup>§</sup> | 30 (1.2) |  |  |
| IgG-4 disease<sup>§</sup> | 30 (1.2) |  |  |
| Myositis<sup>§</sup> | 26 (1.0) |  |  |
| Juvenile idiopathic arthritis<sup>§</sup> | 24 (0.9) |  |  |
| Demyelinating disease<sup>§</sup> | 13 (0.5) |  |  |
| Dominant treatment at time of sampling<sup>+</sup>, n (%)<sup>*</sup> |  |  |  |
| Cytokine inhibitors | 1467 (39.0) |  |  |
| No immunomodulation | 717 (19.1) |  |  |
| Conventional immune modulators | 638 (17.0) |  |  |
| Glucocorticoids | 288 (7.7) |  |  |
| Signaling inhibitors | 239 (6.4) |  |  |
| B-Cell inhibitors | 197 (5.2) |  |  |
| T-Cell inhibitors | 62 (1.6) |  |  |
| Adhesion molecule inhibitors | 45 (1.2) |  |  |
| Other drugs | 106 (2.8) |  |  |
| Combination treatment | 482 (12.8) |  |  |
| Overall glucocorticoid use | 814 (21.7) |  |  |

IMID, immune mediated inflammatory disease; SD, standard deviation; IQR, interquartile range
<sup>§</sup> Designated as “other diagnoses” throughout the manuscript.
<sup>+</sup> Detailed list of individual agents per class provided in Suppl. Table-1
<sup>\*</sup> Counts and percentages based on number of samples.

### Time course of SARS-CoV-2 IgG antibody levels after two vaccinations

Observed geometric mean SARS-CoV-2 IgG antibody levels by periods of follow-up after first vaccination are presented in **Table-2A**. Largest OD values were observed in the 8- to 10-week period after vaccination in both groups, which seemed to be attained earlier in HC compared to IMID patients and with higher mean values. Estimated marginal mean antibody levels after adjustment for age and sex (**Table-2B**) were higher in HC compared to IMIDs at all timepoints after 8 weeks, where the peak marginal mean antibody response in HC (12.48; 95%CI 11.52 to 13.52) was more than twice the value estimated for IMID patients (5.71; 95%CI 5.46 to 5.97) showing a large mean difference of 6.98 (95% CI 5.93 to 8.04). At week 40, the difference was less pronounced with a marginal mean antibody level of 3.31 (95% CI 3.01 to 3.64) in HC compared to 2.40 (95% CI 2.00 to 2.88) in IMID patients, still with a small but significant mean difference of 0.91 (95% CI 0.37 to 1.44). HC showed a biphasic loss of antibodies over time; an initial rapid decline after the peak up to week 20 and a slower decline thereafter until week 40 as opposed to a relatively monophasic loglinear decline after a shallower peak in IMID patients (**Figure-1A**).

**Figure-1.**
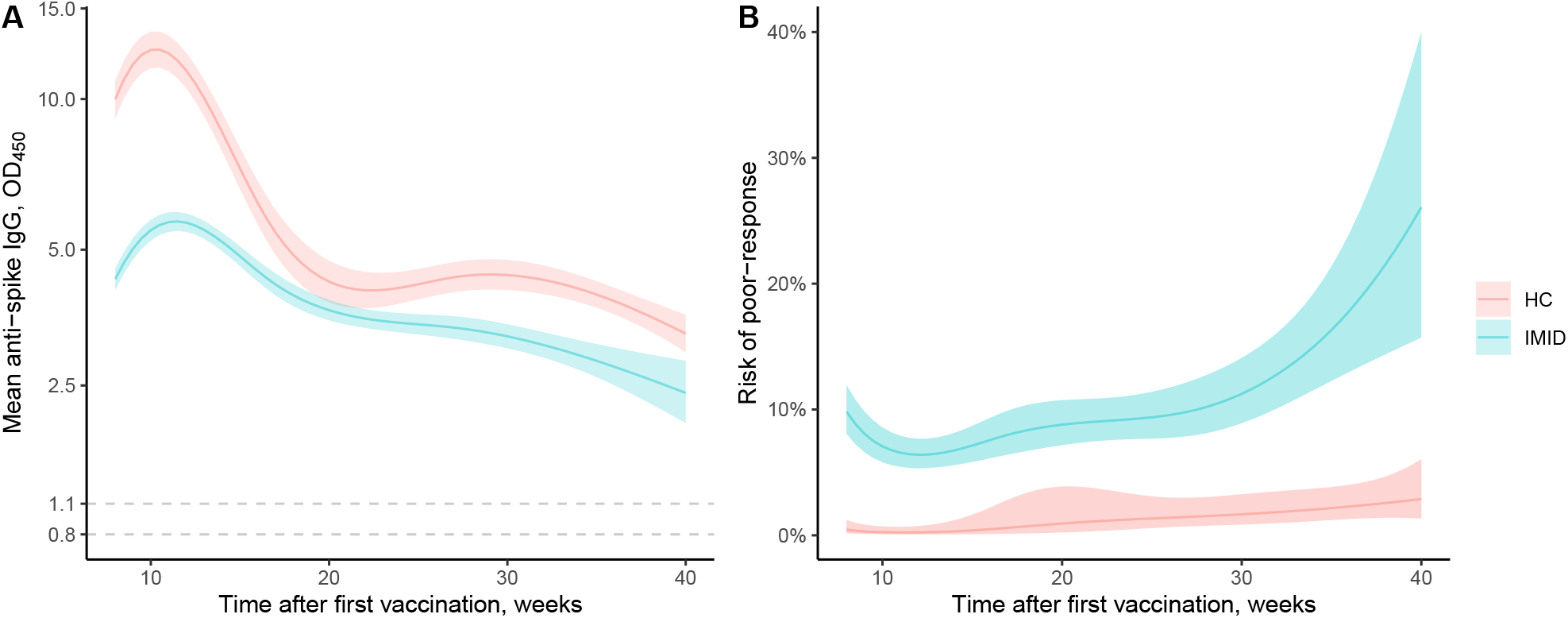
Estimated marginal mean anti-spike IgG levels between 8 to 40 weeks after first vaccination (A). Estimated risks of poor antibody response over time after first vaccination by IMID status. (B) Bands indicate 95% confidence intervals. Dashed lines mark negative (0.8) and borderline (1.1) OD thresholds.

Observed proportion of samples showing poor antibody response (OD<1.1) ranged between 0 to 2.32% among HC, and 7.4% to 17.8% among IMID patients, rising steadily after the 8- to 10-week period (**Table-2A)**. Average estimated marginal risk of poor-response at mean cohort age was higher in IMID patients compared to HC at all timepoints (Figure-1B). These ranged between 0.25% to 2.88% in HC but was above 5% at all time points in IMID patients, reaching 12.82% (95%CI 10.11% to 16.12%) at week 32 and 26.08% (15.71% to 40.03%) at week 40, almost 10 times the risk estimated for HC (2.88%; 95%CI 1.35% to 6.06%) (**Table-2B**).

**Table-2.** Number of samples obtained before 3^rd^ vaccination by post-vaccination period, observed geometric mean optical density and observed number of samples showing poor antibody response (A). Estimated marginal mean antibody levels and estimated risk of poor antibody response with 95% confidence intervals at mean cohort age, at specific timepoints after first vaccination (B). Analyses include samples obtained before 3^rd^ vaccination.

| **A** |  |  |  |  |  |  |
| --- | --- | --- | --- | --- | --- | --- |
| **Period, weeks** | **Healthy controls** |  |  | **IMID patients** |  |  |
|  | **Samples N** | **OD Mean\* (SD)** | **Poor response Samples (%\*)** | **Samples N** | **OD Mean\* (SD)** | **Poor response Samples (%\*)** |
| ≤4 | 168 | 0.29 (4.92) | 131 (77.98) | 266 | 0.43 (5.31) | 175 (65.79) |
| >4 to 8 | 270 | 8.44 (1.56) | 3 (1.11) | 457 | 2.82 (4.04) | 100 (21.88) |
| >8 to 10 | 77 | 8.59 (1.19) | 0 (0) | 297 | 5.19 (2.58) | 22 (7.41) |
| >10 to 16 | 138 | 7.39 (1.76) | 2 (1.45) | 879 | 4.13 (3.21) | 92 (10.47) |
| >16 to 24 | 207 | 6.10 (1.54) | 1 (0.48) | 867 | 3.46 (2.87) | 106 (12.23) |
| >24 to 32 | 417 | 5.08 (1.67) | 5 (1.2) | 637 | 2.98 (2.53) | 85 (13.34) |
| >32 | 431 | 3.87 (1.78) | 10 (2.32) | 174 | 2.72 (2.77) | 31 (17.82) |

| **B** |  |  |  |  |
| --- | --- | --- | --- | --- |
| **Week** | **Healthy controls** |  | **IMID patients** |  |
|  | **OD Mean (95% CI)** | **Poor response % (95% CI)** | **OD Mean (95% CI)** | **Poor response % (95% CI)** |
| 8 | 9.97 (9.17 to 10.84) | 0.45 (0.16 to 1.21) | 4.38 (4.16 to 4.61) | 9.83 (8.06 to 11.94) |
| 10 | 12.48 (11.51 to 13.54) | 0.25 (0.09 to 0.70) | 5.55 (5.28 to 5.82) | 7.07 (5.80 to 8.58) |
| 16 | 6.28 (5.68 to 6.94) | 0.48 (0.10 to 2.21) | 4.56 (4.34 to 4.80) | 7.58 (6.12 to 9.34) |
| 24 | 4.16 (3.85 to 4.49) | 1.27 (0.49 to 3.27) | 3.50 (3.34 to 3.68) | 9.25 (7.64 to 11.15) |
| 32 | 4.34 (4.04 to 4.65) | 1.84 (0.96 to 3.50) | 3.13 (2.93 to 3.35) | 12.82 (10.11 to 16.12) |
| 40 | 3.31 (3.01 to 3.64) | 2.88 (1.35 to 6.06) | 2.40 (2.00 to 2.88) | 26.08 (15.71 to 40.03) |

IMID, immune mediated inflammatory disease; OD optic density, SD, standard deviation, CI, confidence interval.
\* Means and percentages do not take into account multiple samples from singular participants that are expected to be correlated.

Age and gender also had an important impact on the risk of poor response to vaccination. The estimated marginal risk of poor response at week 40 for IMID patients was 17.87% (95%CI 10.12% to 29.59%) at 35 years of age and 35.83% (95%CI 22.74 to 51.43) at 65 years of age, corresponding to an overall odds ratio of 1.03 (95%CI 1.02 to 1.04) per year increase in age across all time points. Male participants were more likely to lose response at week 40, showing an estimated risk of poor response at 28.62% (17.23% to 43.56%) in male IMID patients compared to 23.69 (14.05 to 37.09) in females, corresponding to an overall odds ratio of 1.29 (95% CI 1.02 to 1.63) across all time points.

### SARS-CoV-2 IgG antibody levels by diagnosis

Age and sex-adjusted marginal mean antibody levels by diagnosis showed an overall blunted peak immune response in IMID patients compared to HC (**Suppl. Table-2A, Figure-2**). Of note, peak responses were also lower in untreated IMID patients than in HC (**Suppl. Table-2B, Figure-2**). Largest adjusted mean differences between HC and IMID diagnosis groups were observed around week 10 ranging between 5.62 (95%CI 2.74 to 8.49) for psoriasis and 8.68 (95%CI 6.73 to 10.64) for “other diagnoses”. Lowest responses were found in patients with rheumatoid arthritis, vasculitis and “other diagnoses”. Within IMID groups, marginal mean autoantibody levels at peak were significantly lower in vasculitis and “other diagnoses” in comparison to those with systemic autoimmune diseases, inflammatory bowel diseases, polymyalgia rheumatica, psoriasis and spondyloarthritis with marginal mean differences ranging between 1.57 (95%CI 0.00 to 3.15) and 2.88 (0.38 to 5.37). The peak marginal mean antibody levels for RA were significantly lower than only that of psoriasis among all IMID groups with a mean difference of 2.60 (95%CI 0.11 to 5.09). At week 40, the mean differences in antibody levels between HC and IMID groups were mostly small and imprecise except for RA with a mean difference of 1.49 (95%CI 0.37 to 2.61) and spondyloarthritis with a mean difference of 1.44 (95%CI 0.25 to 2.62).

**Figure-2.**
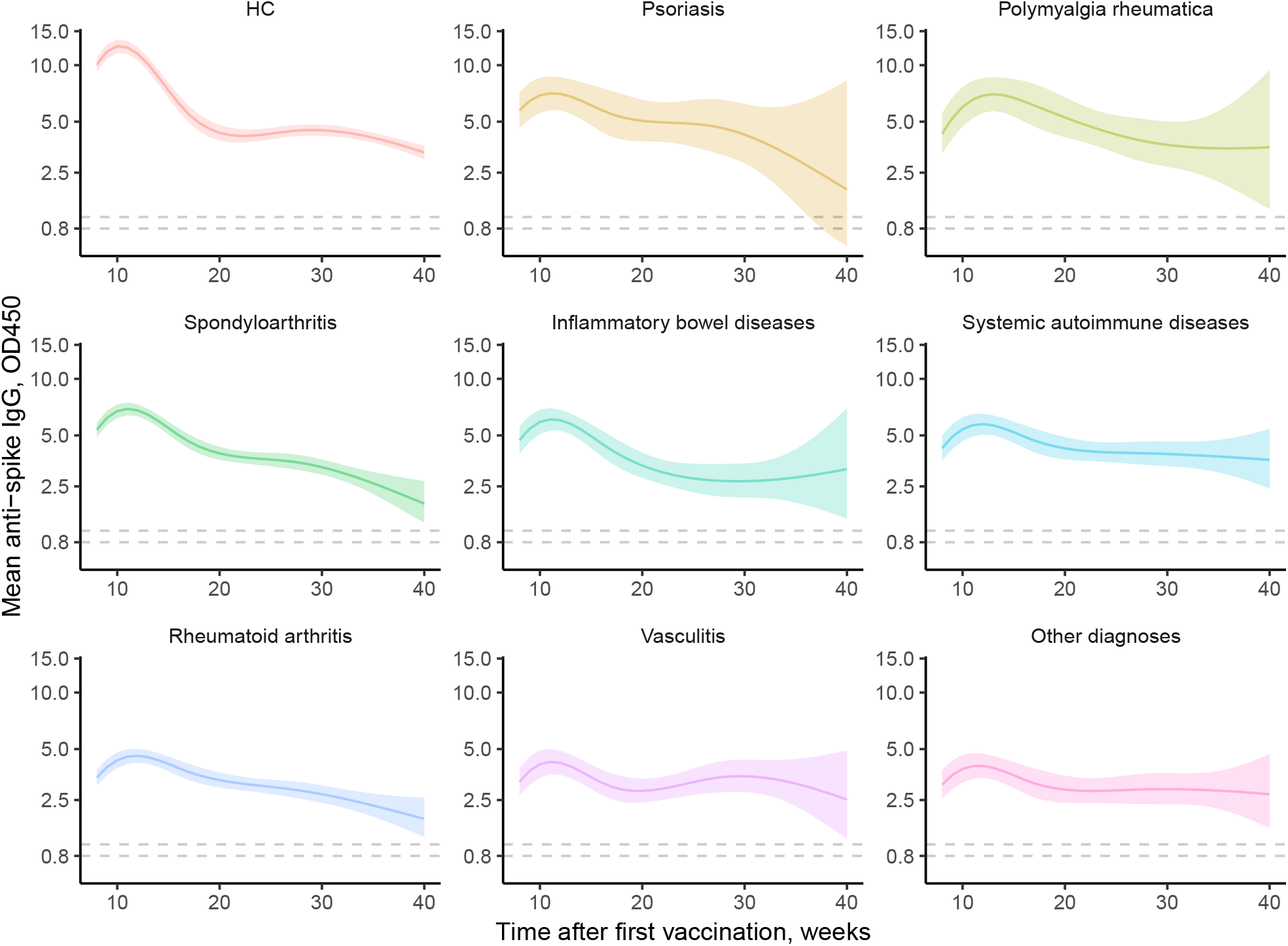
Age and sex adjusted estimated marginal mean anti-spike IgG levels between 8 to 40 weeks after first vaccination by IMID diagnoses. Bands indicate 95% confidence intervals. Panels ordered by peak mean value. Other diagnoses include sarcoidosis, autoinflammatory diseases, IgG-4 disease, inflammatory myopathy, juvenile idiopathic arthritis and inflammatory demyelinating diseases.

### SARS-CoV-2 antibody levels by treatment

The overall course of marginal mean antibody levels was not different in untreated IMID patients from those treated with antimalarials or sulfasalazine (“other agents”) or those treated with glucocorticoid monotherapy (mostly low doses) (**Suppl. Table-2B, Figure-3**). In contrast, mean antibody levels were low throughout follow-up in patients treated with T-cell and B-cell inhibitors. At peak, largest mean differences between HC and treatment groups were observed around week 10 and among these the largest was B cell inhibitors (11.68; 95%CI 10.07 to 13.29). In paired comparisons between treatment groups for peak vaccine response, B-cell inhibitors showed lower mean antibody levels in comparison to all other treatment groups except for T-cell inhibitors. The absolute mean OD differences between B-cell inhibitors and other treatments were in the range of 3.76 (95%CI 2.52 to 4.90) for signaling inhibitors and 6.61 (95%CI 3.58 to 9.64) for hydroxychloroquine or sulfasalazine. Peak responses with T-cell inhibitors were also lower than those observed with cytokine and signaling inhibitors as well as conventional immune modulators and “other drugs” ranging between 2.58 (95%CI 0.99 to 4.17) and 5.36 (95%CI 2.13 to 8.59) (**Suppl. Table-2B**).

**Figure-3.**
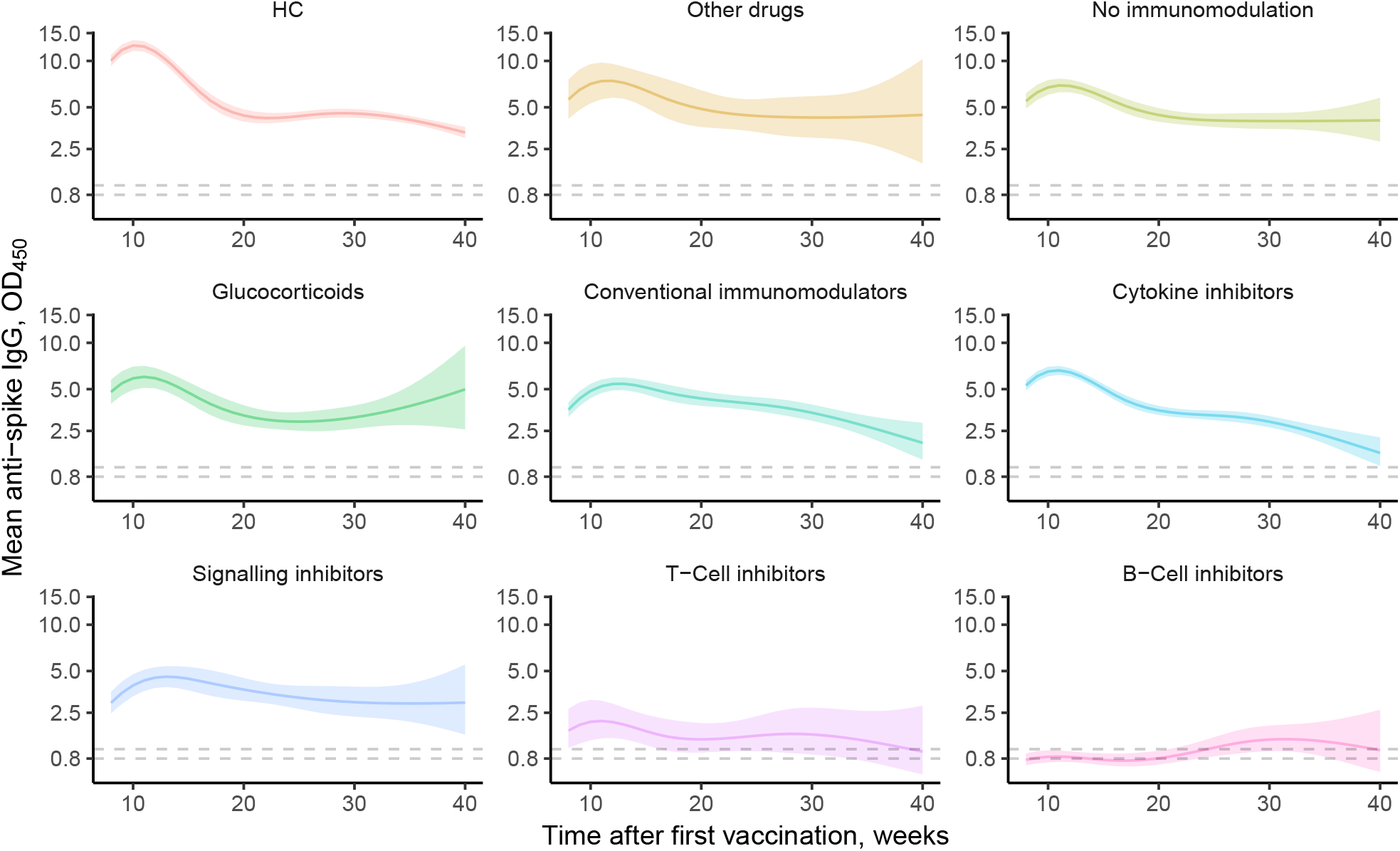
Age and sex adjusted estimated marginal mean anti-spike IgG levels between 8 to 40 weeks after first vaccination by type of treatments used for IMIDs. Bands indicate 95% confidence intervals. Panels are ordered by mean of estimated mean values at all timepoints. Detailed list of individual agents per class provided in Suppl. Table-1.

At week 40, marginal mean antibody levels in HC were significantly higher compared to conventional immune modulators, cytokine inhibitors as well as B-cell and T-cell inhibitors. Adjusted absolute mean differences ranged between 1.35 (95%CI 0.04 to 2.67) for conventional immune modulators and 2.32 (95%CI 0.50 to 4.13) for T cell inhibitors. Other pairwise comparisons between HC and IMID treatment groups were unremarkable. Among IMID treatment groups, untreated IMID patients showed higher mean antibody levels compared to cytokine inhibitors (mean difference 2.48; 95%CI 0.01 to 4.94), B cell inhibitors (3.00; 95%CI 0.18 to 5.83) and T-cell inhibitors (3.05; 95%CI 0.15 to 5.95). While responses in T-cell and B-cell inhibitors were low throughout the observation period, responses in cytokine inhibitors were gradually lost over time.

When combination treatment status was added to the model, monotherapy with cytokine inhibitors, T cell inhibitors and conventional immune modulators was associated with higher mean antibody levels than combination therapy. This was most clearly observed with cytokine inhibitors, early in the time course after vaccination and up to 30 weeks. (**Figure-S2**).

### Time course of SARS-CoV-2 antibodies with two and three vaccinations

Overall 241 (6.5%) participants had a third vaccine (Table-1). **Figure-4** depicts the time course of age and sex adjusted marginal mean antibody levels among IMID patients and HC by overall number of doses received. These reflect the selective application of a third vaccine dose in patients with IMIDs who showed an early poor response to double vaccination. Over the long term. i.e. at 40 weeks, marginal mean antibody levels in IMID patients who received a third dose (4.68. 95%CI 3.80 to 5.76) was higher than those in IMID patients who received 2 doses (2.57, 95%CI 2.13 to 3.09) and also HC who received 2 doses (3.34, 95%CI 3.03 to 3.68).

**Figure-4.**
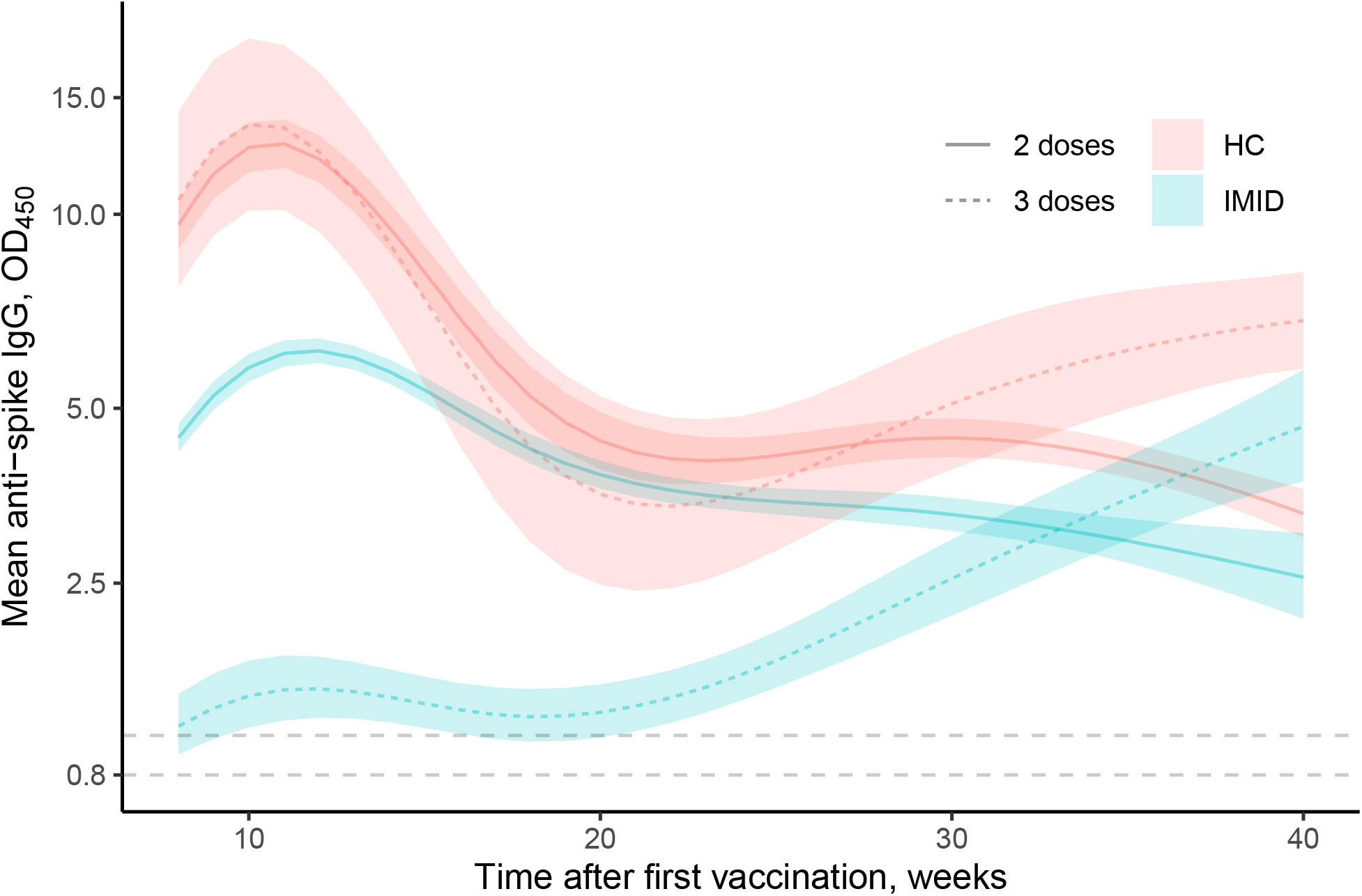
Age and sex adjusted estimated marginal mean anti-spike antibody levels in healthy controls and IMID patients receiving 2 vs 3 doses. Bands indicate 95% confidence intervals.

## Discussion

This large prospective cohort study shows lower intensity and reduced longevity of the antibody response to two doses of SARS-CoV-2 vaccination in IMID patients. Peak response in IMID patients was less than half of that observed in HC after adjustment. Furthermore, IMID patients showed lower mean antibody levels over the entire course of follow-up losing humoral immunity more frequently. Of note, one fifth of younger and one third of older IMID patients lose humoral response 40 weeks after basal immunization. Our results also provide useful insights on the course of vaccine response across various treatment groups. They show particularly poor responses in patients treated with B-cell and T-cell inhibitors. Furthermore, while IMID patients treated with cytokine inhibitors showed peak responses comparable to untreated IMID patients, durability of response was not as good and fell beneath that of untreated IMID patients over time.

To ensure sufficient protection, earlier administration of booster doses may be critical in IMID patients. We had previously shown that IMID patients who did not develop antibodies after two doses of SARS-Cov2 vaccination were likely to develop antibodies after a third dose (16). In this study, we demonstrate that, after the booster, IMID patients could achieve anti-SARS-CoV-2 IgG levels higher than non-boosted HC, even though the initial mean antibody levels after the first two vaccinations were lower. In addition, given the observation that peak antibody levels at 10 weeks after the first vaccination in IMID patients are similar to 16- to 24-week levels in HC, it might be reasonable to administer the third dose within the three to four months of the first dose.

Our study has some limitations. We have collected serum samples in an unscheduled timeline along the routine care of IMID patients, this may have resulted in more frequent or diligent sampling in IMID patients who get to visit the clinic more frequently. To mitigate this potential bias, we have used regression methods that account for within patient correlations using random effects. Samples from IMID patients were obtained predominantly under cytokine blocking treatments, therefore the overall between-group comparisons should reflect the weight of this treatment class, which affects the generalizability of our results. On the other hand, we also observe similar differences between IMID patients not receiving any immunomodulating treatment and HC. We did not attempt to separate the effects of treatments independent of diseases or antibody response across disease activity states, therefore the antibody levels reported here could be considered as average effects of treatments combined with the variety of diseases they are used for or similarly as average effects of diagnoses combined with their routine treatment patterns. We did not collect data on corticosteroid doses, however higher dosed corticosteroid treatments are usually given in combination with cytotoxic agents in conditions such as lupus nephritis or ANCA associated vasculitis. Therefore, their effects could be considered as embedded within those treatment categories and participants using only corticosteroid treatment should be more likely on lower maintenance doses, which may explain the relatively favorable antibody course with corticosteroids in comparison to cytotoxic or anti-cytokine treatments.

In conclusion, these data support the concept that IMID patients are at risk to lose protective humoral response after basic SARS-CoV-2 vaccination, a phenomenon, which has also been observed after SARS-CoV-2 infection (3, 9). Considering that anti-SARS-CoV-2 IgG are a good surrogate of protection against COVID-19 (17), lower overall antibody titers may also reflect a higher susceptibility to breakthrough infections. Hence adapted vaccination schedules that include earlier boosters should be implemented to more adequately protect IMID patients. We hope that these findings will be of use to policymakers responsible for disease control and prevention to reconsider current recommendations.

## Data Availability

All data produced in the present work are contained in the manuscript

## Data Availability

All data produced in the present work are contained in the manuscript

## Data Availability

All data produced in the present work are contained in the manuscript

## Data Availability

All data produced in the present work are contained in the manuscript

## Footnotes

### Contributors

Sample collection: DS, FF, AK, CM, PD, TO, JM, JT, JK, SK, AML, VS, DB, FH, ML AR, MP, FS, MR, SK, AJH, KM, BM, RA; Study design: KT, DS, GS Experiments and data analysis: KT, GK, MYM; Tables and figure: DS, KT and GS. Data interpretation: KT, DS, CB, MS, AK, MFN, GS. Writing of the manuscript: KT, DS, FF, MFN, GS. Critical proof reading of the manuscript: all authors.

### Funding

This study was supported by the Deutsche Forschungsgemeinschaft (DFG-FOR2886 PANDORA and the CRC1181 Checkpoints for Resolution of Inflammation). Additional funding was received by the Bundesministerium für Bildung und Forschung (BMBF; project MASCARA), the ERC Synergy grant 4D Nanoscope, the IMI funded project RTCure, the Emerging Fields Initiative MIRACLE of the Friedrich-Alexander-Universität Erlangen-Nürnberg, and the Else Kröner-Memorial Scholarship (DS, no. 2019_EKMS.27).

### Competing interests

None to declare

### Patient consent for data and publication

Based on ethics approval #157_20 B and the TARDA database.

### Ethics approval

Ethical approval (#157_20 B) to conduct this study was granted by the Institutional Review Board of the University Hospital Erlangen. Written informed consent was obtained from the study participants.

### Data availability statement

All data relevant to the study are included in the article or uploaded as supplementary information.

## Key messages

### What is already known about this subject?

While it is known that SARS-CoV-2 vaccination is effective in the general population, few data on the sustained efficacy of the vaccine in patients with immune-mediated inflammatory diseases (IMIDs) exist at the moment. Most importantly, it is not known whether the immune response against the SARS-CoV-2 vaccine is durable and how it is influenced by immune-modulatory therapy.

### What does this study add?

The study shows that the antibody response of IMID patients after two doses of SARS-CoV-2 vaccination does not only have a lower intensity but also a reduced durability. Especially many older IMID patients lose their vaccination response over time and are likely to profit from booster vaccination. Among immune-modulatory therapies, T cell and B cell targeted agents show the strongest inhibitory effect on the immune response against the SARS-CoV-2 vaccine.

### How might this impact on clinical practice or future developments?

Our data support the introduction of adjusted vaccination schedules with more frequent booster vaccinations in IMID patients to ensure effective immunization to prevent breakthrough infections.

## Notes

### Competing Interest Statement

The authors have declared no competing interest.

### Author Declarations

The study was approved by the Institutional Review Board of Erlangen University Hospital (#157_20 B).

