## Supplementary material for "Intensity and longevity of SARS-CoV-2 vaccination response and efficacy of adjusted vaccination schedules in patients with immune-mediated inflammatory disease": Suppl.

| Suppl. Table-1 Treatment classification. | |
| --- | --- |
| Treatment category | Agent |
| Adhesion molecule inhibitors | Vedolizumab |
|  | Etrolizumab |
| B-Cell inhibitors | Belimumab |
|  | Ocrelizumab |
|  | Rituximab |
| Cytokine inhibitors | Anakinra |
|  | Canakinumab |
|  | Ustekinumab |
|  | Brodalumab |
|  | Ixekizumab |
|  | Secukinumab |
|  | Guselkumab |
|  | Tildrakizumab |
|  | Risankizumab |
|  | Spesolimab |
|  | Dupilumab |
|  | Benralizumab |
|  | Mepolizumab |
|  | Sarilumab |
|  | Tocilizumab |
|  | Adalimumab |
|  | Certolizumab |
|  | Etanercept |
|  | Golimumab |
|  | Infliximab |
| Conventional immune modulators | Azathioprin |
|  | Cyclophosphamide |
|  | Leflunomide |
|  | Methotrexate |
|  | Mycophenolate |
| Other drugs | Chloroquin |
|  | Hydroxychloroquin |
|  | Sulfasalazin |
|  | Mesalazin |
| Signaling inhibitors | Baricitinib |
|  | Filgotinib |
|  | Tofacitinib |
|  | Upadacitinib |
|  | Apremilast |
| T-Cell inhibitors | Cyclosporin A |
|  | Abatacept |

| Suppl. Table-2 Vaccines administered in study time window. | | | | | |
| --- | --- | --- | --- | --- | --- |
|  |  |  | Overall | HC | IMID |
| Total vaccine doses administered in study time window | | | 7495 | 2433 | 5062 |
|  | First dose | BioNTech/Pfizer, Comirnaty | 2826 (75.7) | 949 (79.2) | 1877 (74.0) |
|  |  | AstraZeneca COVID-19 Vaccine | 559 (15.0) | 177 (14.8) | 382 (15.1) |
|  |  | Moderna COVID-19 Vaccine | 246 (6.6) | 56 (4.7) | 190 (7.5) |
|  |  | Johnson & Johnson/Janssen | 55 (1.5) | 9 (0.8) | 46 (1.8) |
|  |  | Unknown/No reply | 49 (1.3) | 7 (0.6) | 42 (1.7) |
|  | Second dose | BioNTech/Pfizer, Comirnaty | 2620 (74.5) | 766 (67.1) | 1854 (78.0) |
|  |  | AstraZeneca COVID-19 Vaccine | 229 (6.5) | 67 (5.9) | 162 (6.8) |
|  |  | Moderna COVID-19 Vaccine | 313 (8.9) | 103 (9.0) | 210 (8.8) |
|  |  | Johnson & Johnson/Janssen | 1 (0.0) | 0 (0.0) | 1 (0.0) |
|  |  | Unknown/No reply | 356 (10.1) | 205 (18.0) | 151 (6.3) |
|  | Third dose | BioNTech/Pfizer, Comirnaty | 195 (80.9) | 92 (97.9) | 103 (70.1) |
|  |  | AstraZeneca COVID-19 Vaccine | 19 (7.9) | 0 (0.0) | 19 (12.9) |
|  |  | Moderna COVID-19 Vaccine | 10 (4.1) | 1 (1.1) | 9 (6.1) |
|  |  | Johnson & Johnson/Janssen | 8 (3.3) | 0 (0.0) | 8 (5.4) |
|  |  | Unknown/No reply | 9 (3.7) | 1 (1.1) | 8 (5.4) |
|  | All doses | BioNTech/Pfizer, Comirnaty | 5641 (75.3) | 1807 (74.3) | 3834 (75.7) |
|  |  | AstraZeneca COVID-19 Vaccine | 807 (10.8) | 244 (10.0) | 563 (11.1) |
|  |  | Moderna COVID-19 Vaccine | 569 (7.6) | 160 (6.6) | 409 (8.1) |
|  |  | Johnson & Johnson/Janssen | 64 (0.9) | 9 (0.4) | 55 (1.1) |
|  |  | Unknown/No reply | 414 (5.5) | 213 (8.8) | 201 (4.0) |

| Suppl. Table-3 Marginal mean antibody levels by IMID diagnosis (A) and treatment (B) at selected time points. | | | | | | | | | |
| --- | --- | --- | --- | --- | --- | --- | --- | --- | --- |
| A | | | | | | | | | |
| Timepoint,  weeks | Healthy Controls | Spondyloarthritis | Rheumatoid arthritis | Systemic autoimmune diseases | Polymyalgia rheumatica | Vasculitis | Inflammatory bowel diseases | Psoriasis | Other |
| 8 | 10.06  (9.25 to 10.93) | 5.36  (4.90 to 5.86) | 3.44  (3.09 to 3.83) | 4.24  (3.61 to 4.98) | 4.28  (3.28 to 5.57) | 3.24  (2.68 to 3.91) | 4.70  (3.95 to 5.58) | 5.77  (4.61 to 7.22) | 3.12  (2.56 to 3.81) |
| 10 | 12.55  (11.57 to 13.61) | 6.77  (6.22 to 7.36) | 4.33  (3.92 to 4.80) | 5.41  (4.65 to 6.29) | 6.01  (4.70 to 7.69) | 4.12  (3.44 to 4.92) | 5.92  (5.08 to 6.90) | 6.93  (5.58 to 8.61) | 3.87  (3.22 to 4.64) |
| 16 | 6.28  (5.68 to 6.95) | 5.01  (4.59 to 5.47) | 3.91  (3.52 to 4.34) | 4.93  (4.26 to 5.69) | 6.44  (5.01 to 8.27) | 3.22  (2.70 to 3.83) | 4.52  (3.82 to 5.33) | 5.79  (4.40 to 7.61) | 3.35  (2.77 to 4.06) |
| 24 | 4.22  (3.90 to 4.55) | 3.69  (3.38 to 4.01) | 3.07  (2.79 to 3.39) | 4.02  (3.54 to 4.57) | 4.41  (3.49 to 5.57) | 3.13  (2.63 to 3.72) | 2.87  (2.37 to 3.48) | 4.90  (3.72 to 6.45) | 2.86  (2.39 to 3.44) |
| 32 | 4.38  (4.08 to 4.69) | 3.04  (2.72 to 3.41) | 2.56  (2.23 to 2.94) | 3.89  (3.27 to 4.62) | 3.60  (2.56 to 5.04) | 3.43  (2.70 to 4.35) | 2.73  (2.08 to 3.58) | 3.79  (2.38 to 6.00) | 2.92  (2.33 to 3.65) |
| 40 | 3.33  (3.03 to 3.67) | 1.90  (1.32 to 2.70) | 1.85  (1.31 to 2.59) | 3.64  (2.42 to 5.44) | 3.59  (1.32 to 9.45) | 2.51  (1.26 to 4.91) | 3.21  (1.44 to 7.02) | 1.91  (0.38 to 8.35) | 2.73  (1.56 to 4.70) |
| B | | | | | | | | | |
| Timepoint, weeks \| Healthy Controls \| Other treatments \| No immunomodulation \| Cytokine inhibitors \| Glucocorticoids \| Conventional immunomodulators \| Signalling inhibitors \| T-Cell inhibitors \| B-Cell inhibitors; 8 \| 10.04 (9.26 to 10.89) \| 5.63 (4.17 to 7.60) \| 5.50 (4.89 to 6.18) \| 5.29 (4.90 to 5.71) \| 4.78 (3.97 to 5.77) \| 3.61 (3.20 to 4.07) \| 2.98 (2.47 to 3.59) \| 1.75 (1.13 to 2.69) \| 0.76 (0.60 to 0.97); 10 \| 12.53 (11.59 to 13.55) \| 7.13 (5.31 to 9.56) \| 6.76 (6.06 to 7.55) \| 6.54 (6.09 to 7.03) \| 5.90 (4.94 to 7.04) \| 4.86 (4.35 to 5.43) \| 3.99 (3.33 to 4.76) \| 2.11 (1.40 to 3.14) \| 0.85 (0.68 to 1.06); 16 \| 6.33 (5.75 to 6.97) \| 6.02 (4.53 to 7.99) \| 5.45 (4.90 to 6.07) \| 4.57 (4.24 to 4.93) \| 4.32 (3.63 to 5.13) \| 4.92 (4.37 to 5.54) \| 4.29 (3.52 to 5.22) \| 1.61 (1.13 to 2.30) \| 0.74 (0.57 to 0.96); 24 \| 4.28 (3.98 to 4.61) \| 4.42 (3.45 to 5.66) \| 4.10 (3.71 to 4.53) \| 3.31 (3.07 to 3.57) \| 2.96 (2.54 to 3.45) \| 4.02 (3.61 to 4.47) \| 3.33 (2.78 to 3.99) \| 1.53 (1.04 to 2.23) \| 1.07 (0.82 to 1.39); 32 \| 4.43 (4.15 to 4.73) \| 4.26 (3.00 to 6.03) \| 4.03 (3.52 to 4.60) \| 2.70 (2.43 to 3.01) \| 3.38 (2.71 to 4.21) \| 3.15 (2.73 to 3.63) \| 2.97 (2.26 to 3.90) \| 1.54 (0.87 to 2.69) \| 1.44 (1.02 to 2.00); 40 \| 3.34 (3.04 to 3.66) \| 4.44 (1.89 to 10.28) \| 4.07 (2.86 to 5.79) \| 1.60 (1.14 to 2.21) \| 4.98 (2.57 to 9.58) \| 1.99 (1.35 to 2.89) \| 3.00 (1.61 to 5.52) \| 1.02 (0.33 to 2.85) \| 1.07 (0.40 to 2.64) | | | | | | | | | |

Controls

N = 1198

Samples = 1805

Total cohort

N=5076

Samples= 11029

Participants with available samples between -4 week of first vaccination and 01/12/2021

N = 5046

Samples = 7289

IMIDs

N= 2535

Samples = 3710

Samples not in study time frame: 3740

Excluded participants: 1313

- Working diagnoses (i.e. investigated for IMIDs with no final diagnosis): 470
- IMID status not ascertained: 368
- Other immune-mediated diseases (e.g. Urticaria, asthma, atopic dermatitis, unclassified arthritis, granulomatous mastitis, uveitis): 278
- Not certainly classifiable as IMID (e.g. Raynaud’s phenomenon, undifferentiated connective tissue disease): 86
- Immunodeficiency: 37
- Organ specific autoimmunity: 32
- Crystal arthritis: 29
- Malignancy: 13

Suppl. Figure-1 Flow-chart describing cohort selection.

Suppl. Figure-2 Marginal mean antibody levels by treatment categories stratified by monotherapy vs combination treatment status.

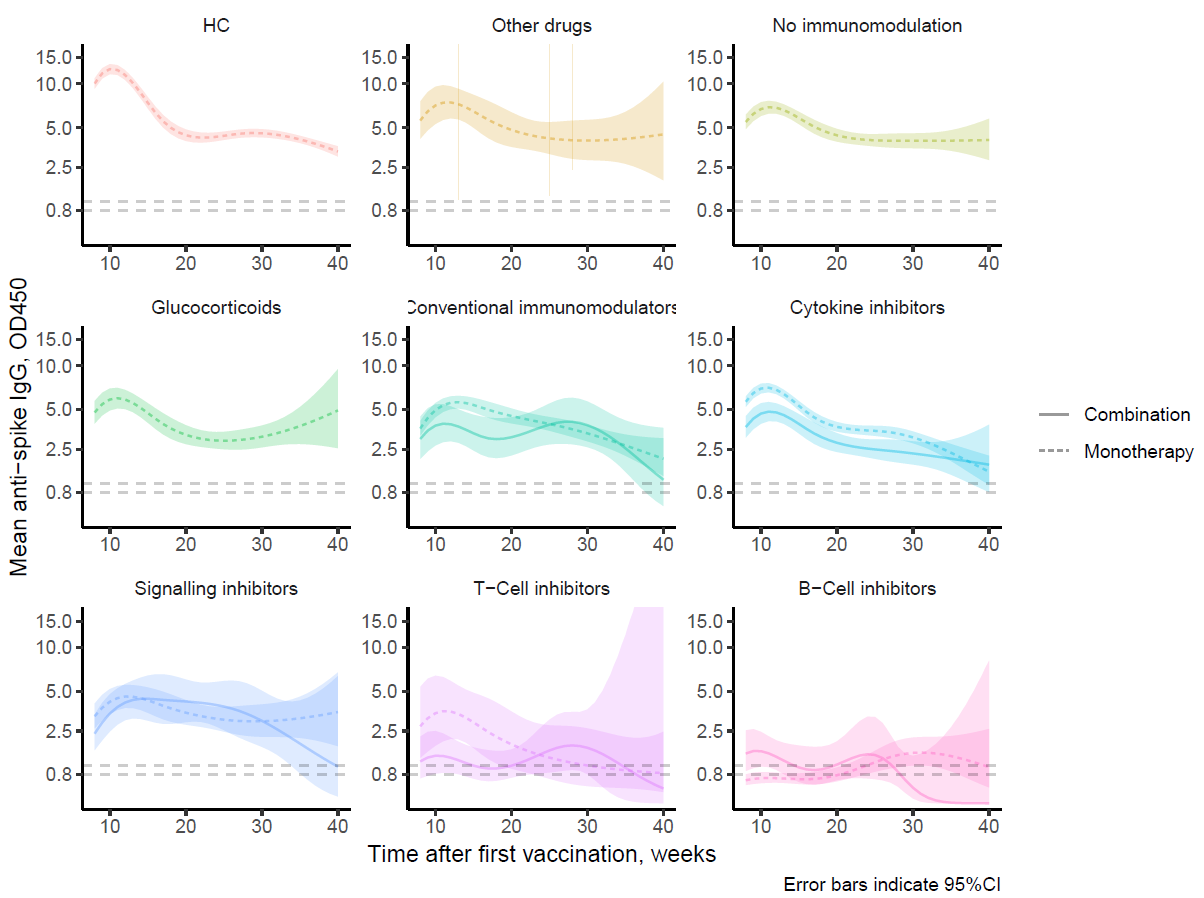
